# Clinical utility of the 4S-AF scheme in predicting recurrence of atrial fibrillation after radiofrequency catheter ablation

**DOI:** 10.1101/2024.07.02.24309867

**Authors:** Naiyuan Cui, Haiwei Li, Weiping Sun, Zefeng Wang, Zhongyu Yuan, Botao Zhu, Yutong Liu, Huanfu Liu, Yongquan Wu, Xiaoping Zhang

## Abstract

**Background:** The 4S-AF scheme, consisting of four domains related to atrial fibrillation (AF) [stroke risk (St), symptoms (Sy), severity of AF burden (Sb), and substrate (Su)], represents a novel approach for the structural characterization of AF. We aimed to assess the clinical utility of the scheme in predicting AF recurrence after radiofrequency catheter ablation (RFCA).

**Methods:** We prospectively enrolled 345 consecutive patients with AF who underwent initial RFCA between January 2019 and December 2019. The 4S-AF scheme score was calculated and used to characterize AF. The primary outcome assessed was AF recurrence after RFCA, defined as any documented atrial tachyarrhythmia episode lasting at least 30 seconds.

**Results:** Of 345 patients [age 61 (IQR: 53-68) years, 34.2% female, 70.7% paroxysmal AF] were analyzed. The median duration of AF history was 12 (IQR: 3-36) months, and the median number of comorbidities was 2 (IQR: 1-3), and 157 (45.5%) patients had left atrial enlargement. During a median follow-up period of 28 (IQR: 13-37) months, AF recurrence occurred in 34.4% of patients. Both 4S-AF scheme score (HR 1.38, 95% CI: 1.19–1.59, P<0.001) and 2S-AF scheme by eliminating the Sy and St domains (HR 1.59, 95% CI: 1.33–1.89, P<0.001) were independent predictors of AF recurrence after RFCA. For each domain, we found that the independent predictors were Sb (HR 1.84, 95% CI: 1.25–2.72, P=0.002) and Su (HR 1.71, 95% CI: 1.36–2.14, P<0.001). Furthermore, 4S-AF scheme score (AUC 65.2%, 95% CI: 59.3–71.1) and 2S-AF scheme score (AUC 66.2%, 95% CI: 60.2–had a modest ability to predict AF recurrence after RFCA.

**Conclusions:** The novel 4S-AF scheme is feasible for evaluating and characterizing AF patients who undergo RFCA and aids us in identifying patients with a high risk of AF recurrence after RFCA.

**CLINICAL PERSPECTIVE:** *What Is New?:*

- The 4S-AF scheme can be used to characterize and assess patients with atrial fibrillation (AF) who will undergo radiofrequency catheter ablation (RFCA).
- A higher 4S-AF scheme score and 2S-AF scheme score modified by eliminating the Sy and St domains of the 4S-AF scheme score were significantly associated with an elevated atrial fibrillation (AF) recurrence rate after RFCA.
- Severity of AF burden (Sb) and substrate (Su) domain scores were independently associated with AF recurrence outcomes after RFCA.
- 4S-AF scheme score and 2S-AF scheme score had modest predictive capability for AF recurrence.

*What Are the Clinical Implications?:*

- AF is a common cardiac arrhythmia and recurrence rates of AF after RFCA are substantial.
- Using the 4S-AF scheme to characterize AF may aid in identifying high-risk patients who need to receive RFCA in terms of AF recurrence.
- Compared to existing risk stratification methods, such as HATCH and CHA_2_DS_2_-VASc score, 4S-AF scheme score and 2S-AF scheme score had better clinical predictive capability of AF recurrence.

## Introduction

Atrial fibrillation (AF) is a heterogeneous and complex cardiac arrhythmia with a steadily increasing global incidence and prevalence. AF can severely impair patients’ quality of life due to its serious complications, including stroke, cognitive impairment, heart failure (HF), and sudden cardiac death.^1–3^ Numerous studies have confirmed the effectiveness of radiofrequency catheter ablation (RFCA) in maintaining sinus rhythm and reducing the burden of AF, thereby improving cardiovascular outcomes.^4–7^ RFCA is currently the first-line rhythm control therapy and the cornerstone of AF treatments in patients with symptomatic, drug-resistant AF.^5,8^

However, recurrence rates of AF after RFCA are substantial and are estimated to range from 20 to 75% two years after the initial RFCA.^9^ The recurrence of AF can be attributed to technical failure, extrapulmonary vein triggers, autonomic neural activity, and AF progression.^8–11^ Evidence from both animal models and clinical studies indicates that atrial cardiomyopathy based on atrial fibrosis and atrial remodelling may be an underlying arrhythmia substrate that plays a pivotal role in AF progression and AF recurrence after RFCA.^12–14^ In addition, major cardiovascular (CV) risk factors for AF, such as aging, obesity, hypertension, diabetes mellitus (DM), HF, and coronary artery disease (CAD), promote atrial electrical and structural remodelling leading to AF progression and AF recurrence.^1,15–19^ Therefore, there is an urgent need for simple tools and integrated models to assess the complex arrhythmia mechanism and identify individuals who are likely to benefit from RFCA in routine clinical practice.

Recently, Potpara TS et al. proposed a novel structured characterization scheme for AF called 4S-AF scheme which consists of four domains: stroke risk (St), symptom severity (Sy), severity of AF burden (Sb), and substrate (Su).^20^ Emerging studies have shown that this novel scheme could provide prognostic information on AF progression and its adverse outcomes.^21–26^ As this novel structured pathophysiology-based characterization scheme facilitates AF assessment and therapy decisions, it has been adopted by the 2020 European Society of Cardiology (ESC) AF guidelines for holistic and personalized AF management.^5^ However, its clinical usefulness in AF patients undergoing RFCA remains unknown. Therefore, this study aimed to determine the clinical utility of the 4S-AF scheme in the prediction of AF recurrence after RFCA.

## Methods

### Study design and population

This study is a single-centre, prospective, observational cohort study. The study population comprised 545 consecutive patients with AF who underwent RFCA in Beijing Anzhen Hospital between January 2019 and December 2019. Inclusion criteria were (1) age>18 years; (2) a diagnosis of AF and underwent RFCA treatment;^5^ (3) voluntary participation in this study and signed informed consent. Patients were excluded if they had (1) long-standing persistent and permanent AF (n=102); (2) history of coronary artery bypass graft surgery (CABG) or other cardiac surgeries (n=7); (3) severe renal dysfunction [estimated glomerular filtration rate (eGFR)<30ml/min/1.73m^2^] (n=5); (4) previous RFCA history for AF (n=29). Finally, the study cohort consisted of 402 patients who had signed the informed consent. This study was designed and performed in accordance with the Declaration of Helsinki for Human Research and was approved by the Beijing Anzhen Hospital Ethics Committee (Approval No: 2024103X).

### Data collection

Demographic, clinical, laboratory, and echocardiographic data at baseline were extracted from the medical records. The demographic and clinical data included age, gender, body mass index (BMI), total AF history, AF classifications, EHRA score, comorbidity, and medication history. Two independent cardiologists calculated and determined the CHA2DS2-VASc score and European Heart Rhythm Association (EHRA) symptom score.^5^ The HATCH score was also calculated using 1 point each for hypertension (H), age ≥ 75 years (A), and chronic obstructive pulmonary disease (C), and 2 points each for transient ischemic attack (TIA) or stroke history (T) and HF (H).^27^ Echocardiographic parameters were measured by a Philips 7C color Doppler ultrasound before the operation, including left atrial diameter (LAD), left ventricular end-systolic diameter (LVESD), left ventricular end-diastolic diameter (LVEDD), and left ventricular ejection fraction (LVEF). Left atrial volume (LAV) was evaluated by the prolate ellipse method using the formula: LAD1 (anterior-posterior) × LAD2 (superior-inferior) × LAD3 (medial-lateral) × 0.523.^28^ Body surface area (BSA) was calculated using the Mosteller formula. Left atrial volume index (LAVI) was calculated by LAV/BSA.^29^

### 4S-AF scheme characterization

All patients were characterized based on 4S-AF scheme comprised of four domains: stroke risk (St), symptoms (Sy), severity of AF burden (Sb), and substrate (Su) (Table 1). St was assessed if at non-low stroke risk and had indication for oral anticoagulant therapy based on CHA2DS2-VASc score. Sy ranging from none to disabling were characterized using EHRA symptom score. Sb was defined as the duration and frequency of the AF episodes based on the AF classification of the 2020 ESC guidelines.^5^ Su was characterized based on three subdomains: comorbidity/CV risk factors score, left atrial (LA) enlargement score, and Age>75 score. The number of comorbidities was calculated by following comorbidities: hypertension, HF, hypercholesterolaemia, DM, CAD, peripheral artery disease, obesity (BMI>30 kg/m^2^), kidney dysfunction (eGFR<60ml/min/1.73m^2^) and moderate or severe mitral valve regurgitation.^20–26^ 4S-AF scheme score was the sum of each domain with a maximum score of 10 (St=1, Sy=2, Sb=2, and Su=5). 2S-AF scheme score was modified by eliminating the Sy and St domains of the 4S-AF scheme score. The interpretation and definition of each domain are presented in Table 1.

**Table 1.**
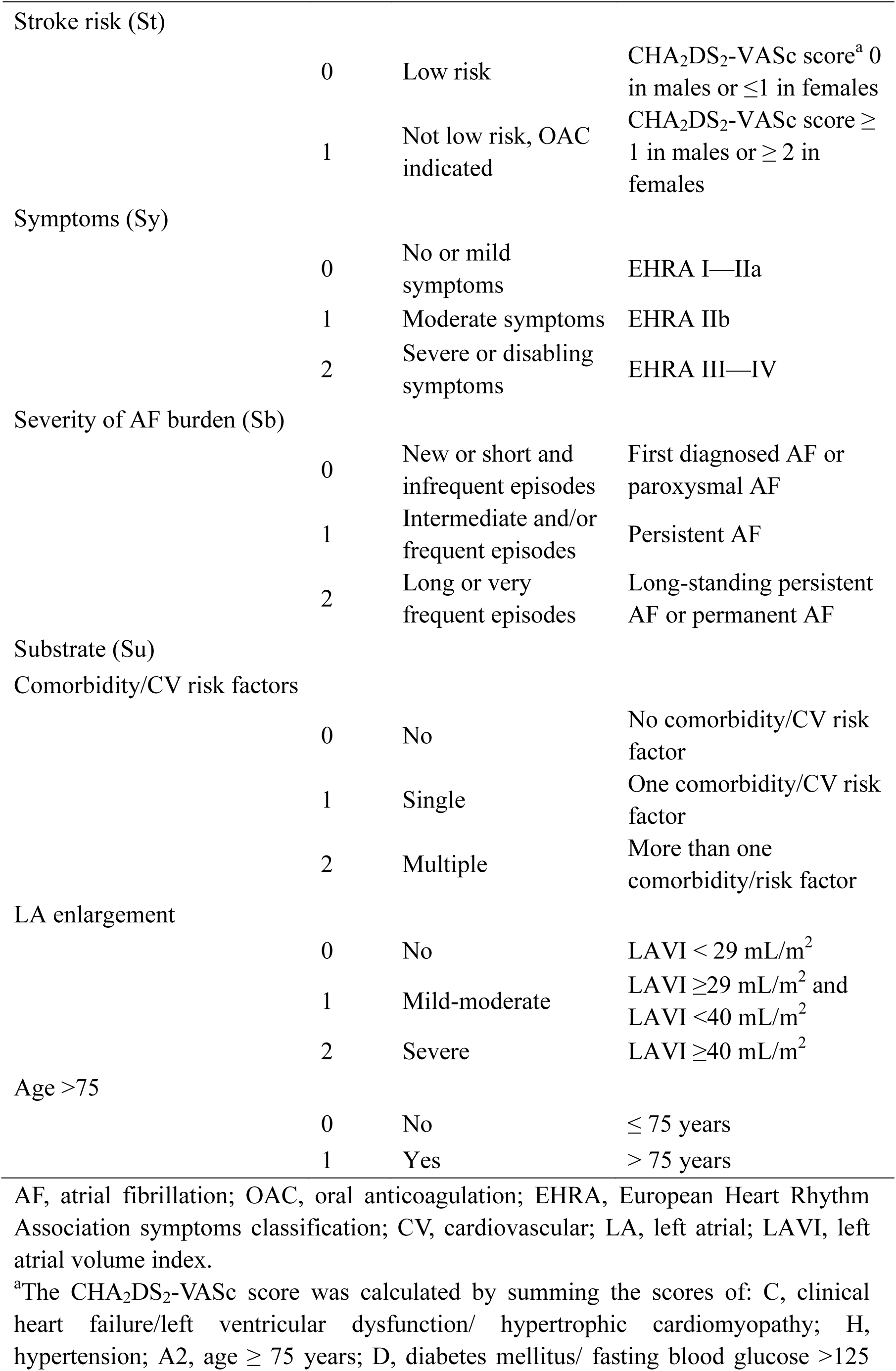
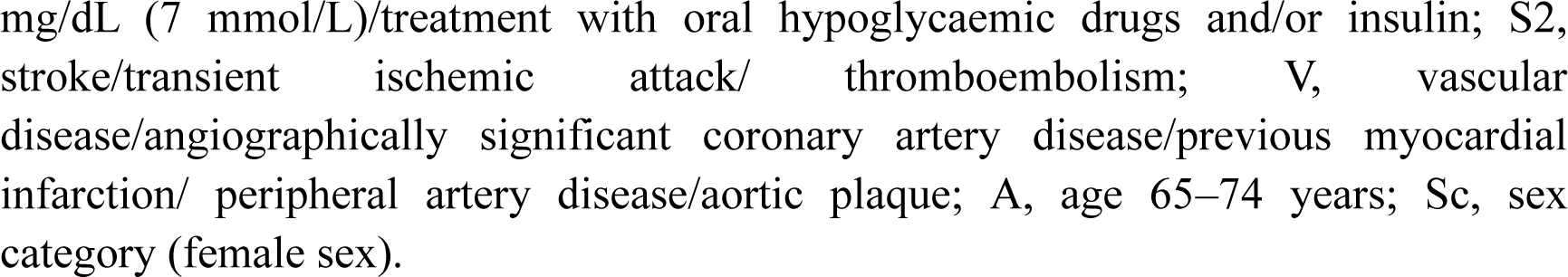
Domains, interpretation and definition of the 4S-AF scheme.

### Outcome and follow-up

Follow-up was performed after ablation by telephone and regular visits to our outpatient clinics. Furthermore, patients were strongly recommended to obtain a 12-lead electrocardiogram (ECG) at the nearest hospital if they experienced any symptoms that could be attributed to arrhythmia or noticed any irregular pulse by routine self-palpitation or auscultation. The outcome was AF recurrence defined as any documented atrial tachyarrhythmia (AF, atrial flutter, or atrial tachycardia) episode lasting for at least 30 seconds after ablation.^11^ To accurately adjudicate outcomes, a detailed medical and physical examination, 12-lead ECG, and 24-hour Holter monitoring were performed at each visit and confirmed by trained study personnel and cardiologists. Follow-up data were based on telephone and outpatient medical records at Beijing Anzhen Hospital.

### Statistical analysis

Continuous variables with normal distribution were expressed as mean ± standard deviation (SD), and non-normally distributed variables were described as the median and interquartile range (IQR). Categorical variables were expressed as observed number with percentage. Continuous variables were compared between groups using independent Student’s t-test or the Mann–Whitney U test, as appropriate. Categorical data were compared using the chi-squared test or Fisher’s exact test, as appropriate. Univariate and multivariate Cox regression analyses were performed by including each domain of the 4S-AF scheme as covariates to determine risk factors for AF recurrence, and the hazard ratio (HR) and 95% CI were calculated. Multivariable Cox regression analysis was performed based on 4S-AF scheme and 2S-AF scheme after adjustment for age, gender, BMI, CAD, DM, HF, hypertension, stroke/transient ischemic attack (TIA), atherosclerosis, and eGFR. Time-dependent survival between groups was evaluated using Kaplan-Meier curves and compared using the log-rank test. The predictive capability of the 4S-AF and 2S-AF scheme for AF recurrence was tested using receiver-operating characteristic (ROC) curves and area under the curve (AUC) analysis. All statistical testing was two-sided with ɑ=0.05 and performed using SPSS version 24 (IBM Corp, Armonk, NY, USA) or GraphPad Prism (GraphPad version 8.0.0, GraphPad Software, San Diego, CA). Two-tailed P-values of < 0.05 were considered statistically significant.

## Results

### Baseline characteristics

A total of 402 patients with AF who underwent initial RFCA between January 2019 and December 2019 were consecutively included. Among the enrolled patients, 38 patients were lost to follow-up, and 19 patients had incomplete echocardiographic information for atrial substrate assessment (Figure 1). The baseline clinical characteristics of the 345 patients with AF included in our analyses are shown in Tables 2 and 3. The median age was 61 (IQR: 53-68) years, 118 (34.2%) patients were female, and the median total duration of AF was 12 (IQR: 3-36) months. The most common CV risk factors were hypertension (61.4%), HF (39.4%), CAD (16.8%), DM (19.1%) and obesity (16.5%). In terms of echocardiographic parameters, the LAD [42.0 (38.0–47.0) vs. 40.0 (38.0–43.2), p < 0.001], LAV [58.6 (46.8–72.0) vs. 50.8 (43.9–61.9), P<0.001] and LAVI [30.8 (26.4–36.4) vs. 27.0 (23.5–32.3), p < 0.001] of the AF recurrence group were greater than those of the non-recurrence group and differed significantly between the two groups. There were no significant differences in other clinical characteristics, including age, BMI, medical history, AF-related score, and medication between the recurrence and non-recurrence groups.

**Figure 1.**
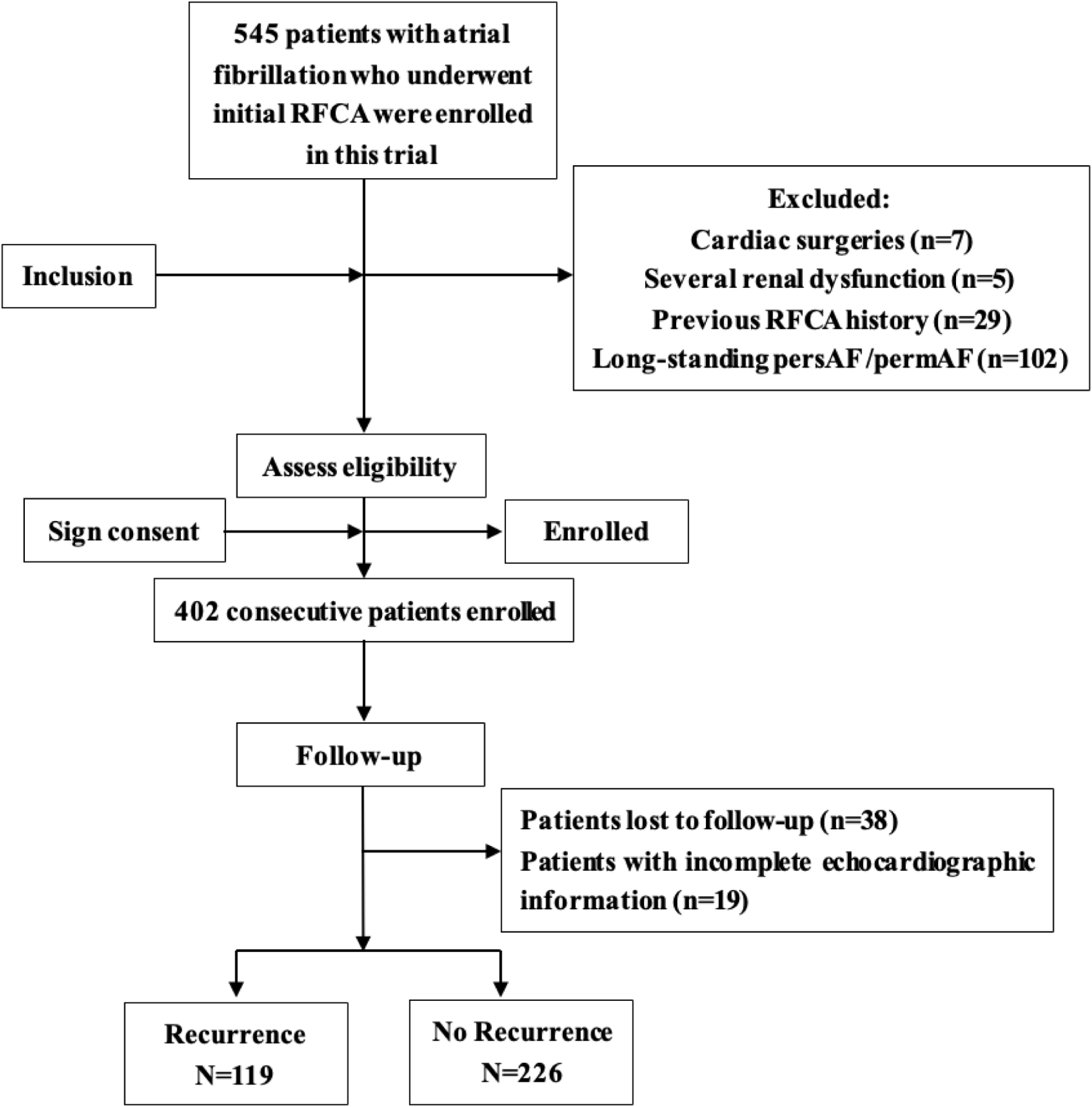
Flow chart of the study. RFCA, radiofrequency catheter ablation; persAF, persistent AF; permAF, permanent AF.

**Table 2.**
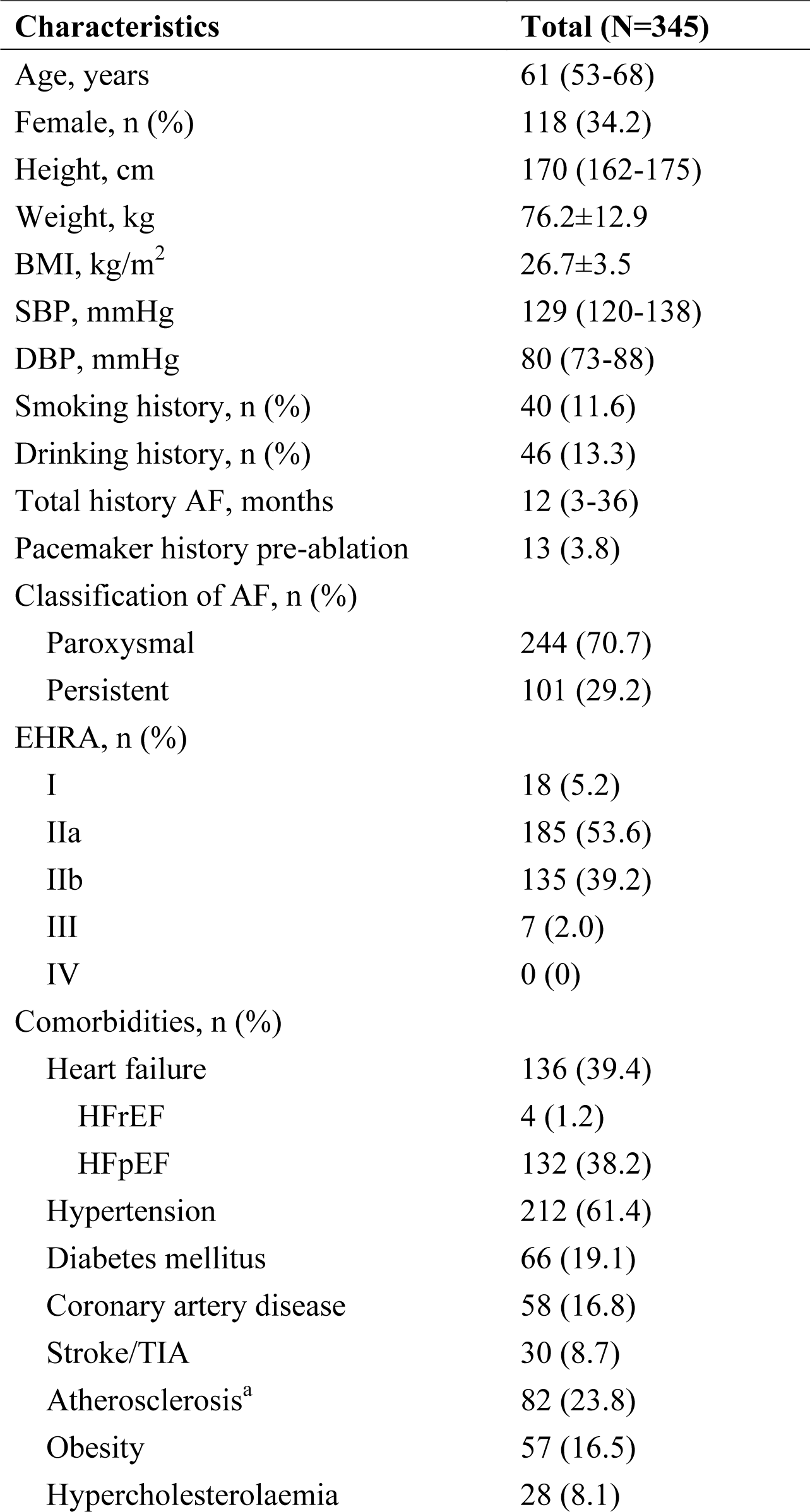
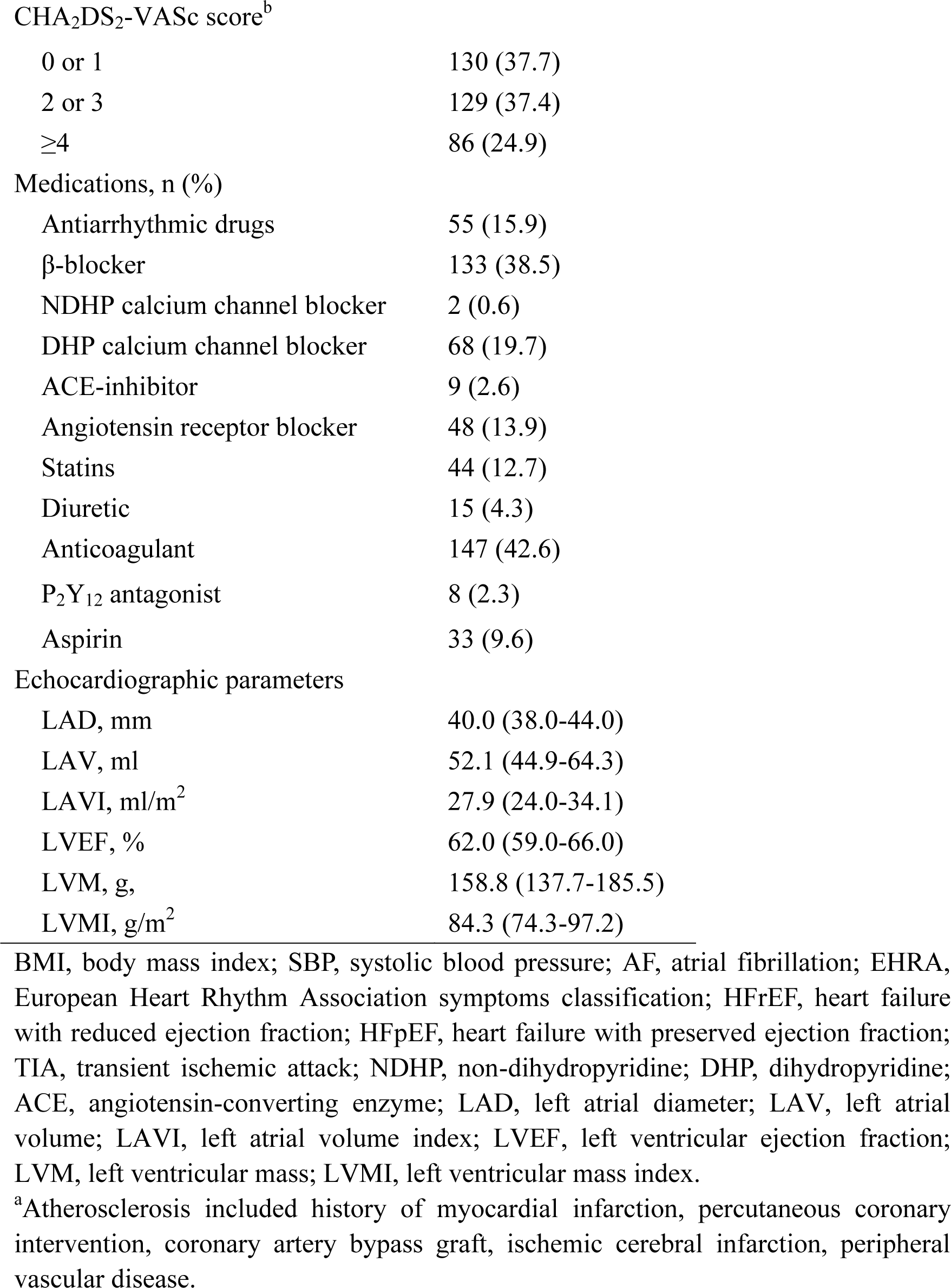
Baseline characteristics.

| Characteristics | Total (N=345) |
| --- | --- |
| Age, years | 61 (53-68) |
| Female, n (%) | 118 (34.2) |
| Height, cm | 170 (162-175) |
| Weight, kg | 76.2±12.9 |
| BMI, kg/m<sup>2</sup> | 26.7±3.5 |
| SBP, mmHg | 129 (120-138) |
| DBP, mmHg | 80 (73-88) |
| Smoking history, n (%) | 40 (11.6) |
| Drinking history, n (%) | 46 (13.3) |
| Total history AF, months | 12 (3-36) |
| Pacemaker history pre-ablation | 13 (3.8) |
| Classification of AF, n (%) |  |
| Paroxysmal | 244 (70.7) |
| Persistent | 101 (29.2) |
| EHRA, n (%) |  |
| I | 18 (5.2) |
| IIa | 185 (53.6) |
| IIb | 135 (39.2) |
| III | 7 (2.0) |
| IV | 0 (0) |
| Comorbidities, n (%) |  |
| Heart failure | 136 (39.4) |
| HFrEF | 4 (1.2) |
| HFpEF | 132 (38.2) |
| Hypertension | 212 (61.4) |
| Diabetes mellitus | 66 (19.1) |
| Coronary artery disease | 58 (16.8) |
| Stroke/TIA | 30 (8.7) |
| Atherosclerosis<sup>a</sup> | 82 (23.8) |
| Obesity | 57 (16.5) |
| Hypercholesterolaemia | 28 (8.1) |
| CHA<sub>2</sub>DS<sub>2</sub>-VASc score<sup>b</sup> |  |
| 0 or 1 | 130 (37.7) |
| 2 or 3 | 129 (37.4) |
| ≥4 | 86 (24.9) |
| Medications, n (%) |  |
| Antiarrhythmic drugs | 55 (15.9) |
| β-blocker | 133 (38.5) |
| NDHP calcium channel blocker | 2 (0.6) |
| DHP calcium channel blocker | 68 (19.7) |
| ACE-inhibitor | 9 (2.6) |
| Angiotensin receptor blocker | 48 (13.9) |
| Statins | 44 (12.7) |
| Diuretic | 15 (4.3) |
| Anticoagulant | 147 (42.6) |
| P<sub>2</sub>Y<sub>12</sub> antagonist | 8 (2.3) |
| Aspirin | 33 (9.6) |
| Echocardiographic parameters |  |
| LAD, mm | 40.0 (38.0-44.0) |
| LAV, ml | 52.1 (44.9-64.3) |
| LAVI, ml/m<sup>2</sup> | 27.9 (24.0-34.1) |
| LVEF, % | 62.0 (59.0-66.0) |
| LVM, g, | 158.8 (137.7-185.5) |
| LVMI, g/m<sup>2</sup> | 84.3 (74.3-97.2) |

BMI, body mass index; SBP, systolic blood pressure; AF, atrial fibrillation; EHRA, European Heart Rhythm Association symptoms classification; HFrEF, heart failure with reduced ejection fraction; HFpEF, heart failure with preserved ejection fraction; TIA, transient ischemic attack; NDHP, non-dihydropyridine; DHP, dihydropyridine; ACE, angiotensin-converting enzyme; LAD, left atrial diameter; LAV, left atrial volume; LAVI, left atrial volume index; LVEF, left ventricular ejection fraction; LVM, left ventricular mass; LVMI, left ventricular mass index.
<sup>a</sup>Atherosclerosis included history of myocardial infarction, percutaneous coronary intervention, coronary artery bypass graft, ischemic cerebral infarction, peripheral vascular disease.

**Table 3.**
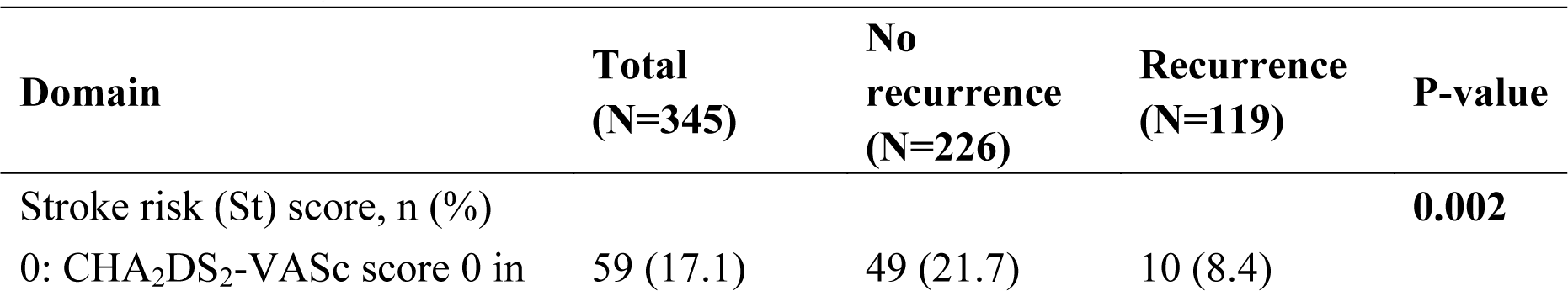
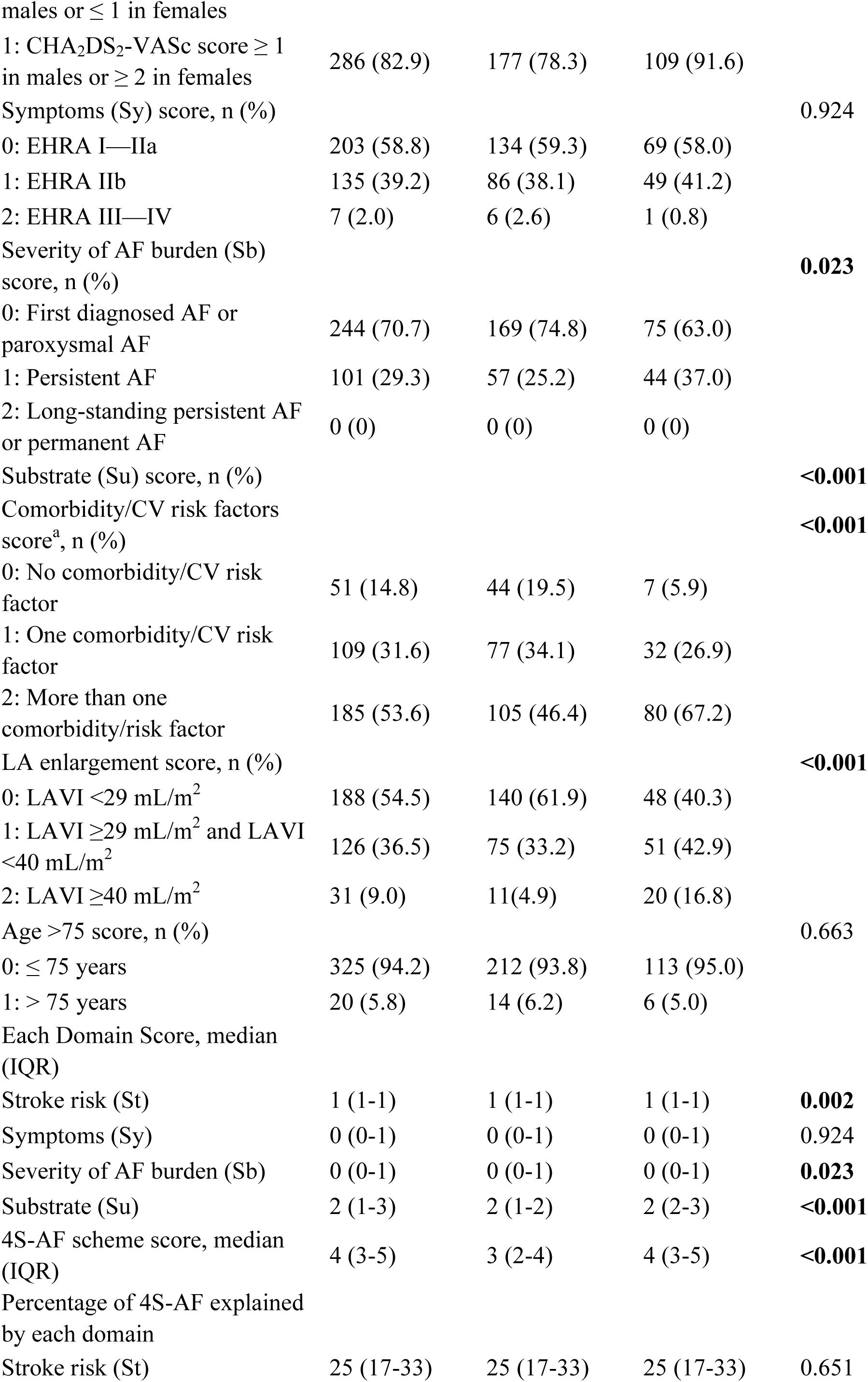
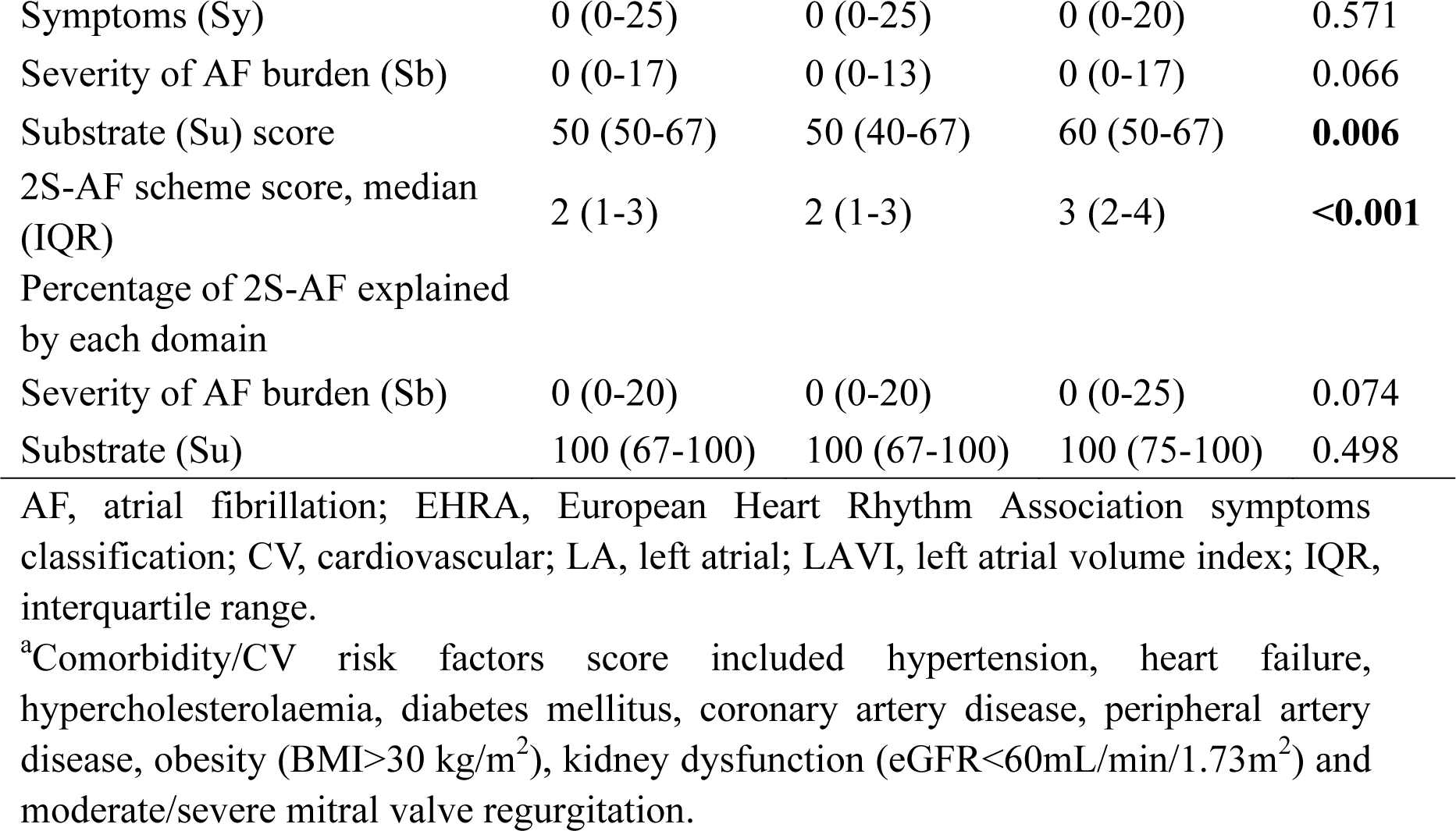
Domains, definition and characterization of 4S-AF scheme.

### Characterization of 4S-AF scheme

Characterization of patients according to the 4S-AF scheme and various domains of the scheme is shown in Table 3. More than half of the patients did not have a low stroke risk and had indications for oral anticoagulant therapy [non-recurrence group, n=177 (78.3%); recurrence group, n =109 (91.6%), P=0.002]. The majority exhibited no or mild symptoms based on the EHRA classification [EHRA I (5.2%), EHRA IIa (53.6%), EHRA IIb (39.2%), EHRA III (2.0%), and EHRA IV (0)]. Additionally, most patients had multiple comorbidities and/or CV risk factors, with a median number of comorbidities was 2 (IQR: 1-3). Furthermore, over half of the patients in the recurrence group had mild to severe LA enlargement [non-recurrence group, n=86 (38.0%); recurrence group, n=71 (59.7%), P<0.001], and more than half of those in the non-recurrence group had no LA enlargement [non-recurrence group, n=140 (61.9%); recurrence group, n=48 (40.3%), P<0.001]. The median score of the 4S-AF scheme was 4 (IQR: 3-5), and for the 2S-AF scheme it was 2 (IQR: 1-3). According to our analysis, the characterization of AF patients using the 4S-AF scheme was significantly explained by the Su domain score, as shown in Table 3 [non-recurrence group, 50 (40-67); recurrence group, 60 (50-67), P=0.006]. However, the percentage of 2S-AF scheme scores explained by each domain was comparable between the two groups.

### Impact of 4S-AF scheme and its domains on AF recurrence outcomes

The median follow-up period was 28 (IQR: 13-37) months after ablation. A total of 119 (34.4%) of the 345 patients included in our analyses experienced AF recurrence after ablation. Using Kaplan‒Meier survival curves, we found that a higher 4S-AF scheme score was significantly associated with an elevated AF recurrence rate (log-rank P<0.001, Figure 2). Moreover, the associations between each domain score and the risk of experiencing AF recurrence outcomes are shown in Figure 3. To investigate the associations between the 4S-AF scheme score and the risk of experiencing AF recurrence, we used univariate and multivariable Cox regression analyses. Univariate Cox regression analysis revealed that the 4S-AF scheme score was an independent predictor of AF recurrence (HR 1.37, 95% CI: 1.22–1.53, P<0.001; Table 4). With adjusted multivariable Cox proportional hazards models, we found that each 1-point increase in the 4S-AF scheme score was significantly associated with a 38% increase in the incidence of AF recurrence (aHR 1.38, 95% CI: 1.19–1.59, P<0.001; Table 4).

**Figure 2.**
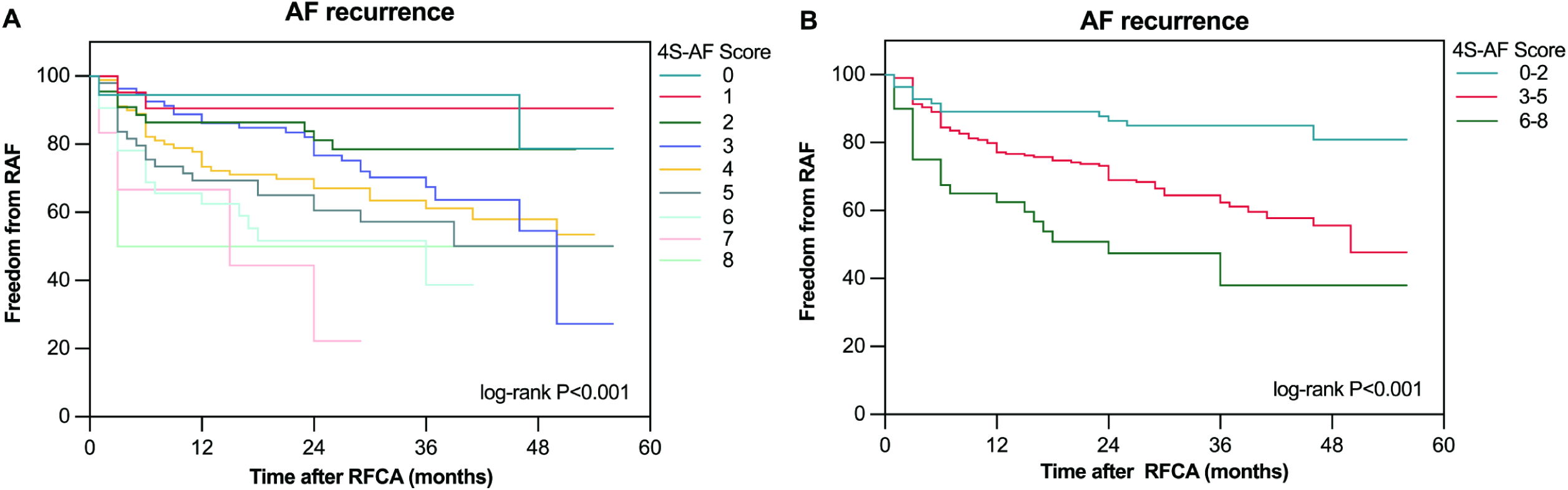
The Kaplan–Meier curve demonstrated the difference in AF recurrence after RFCA between different 4S-AF score. RAF, recurrence of atrial fibrillation; AF, atrial fibrillation; RFCA, radiofrequency catheter ablation.

**Figure 3.**
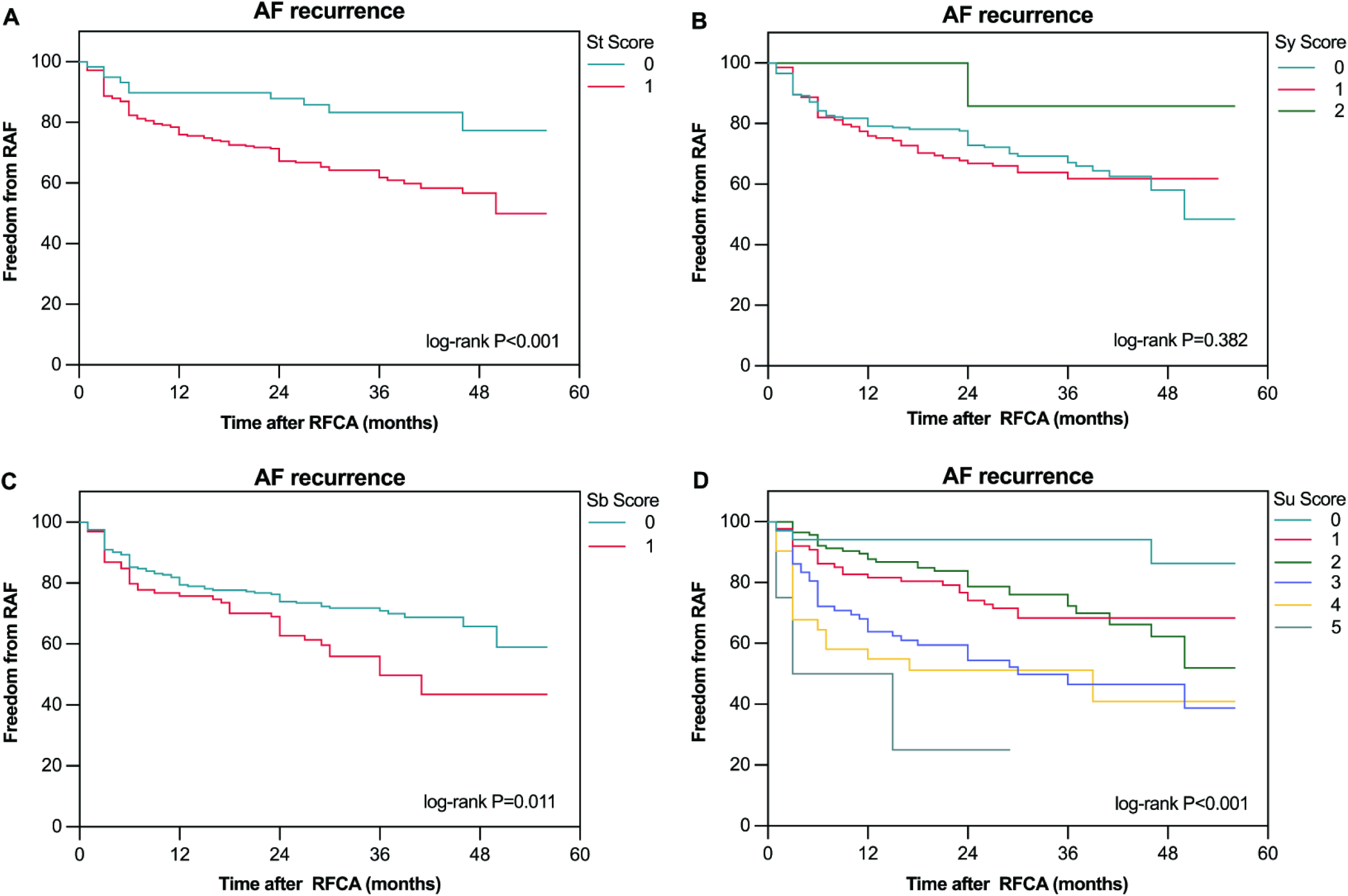
The Kaplan–Meier curve demonstrated the difference in AF recurrence after RFCA between different score of each domain, St domain (A), Sy domain (B), Sb domain (C), Su domain (D). RAF, recurrence of atrial fibrillation; AF, atrial fibrillation; RFCA, radiofrequency catheter ablation; St, stroke risk; Sy, symptoms; Sb, severity of AF burden; Su, substrate.

**Table 4.** Association of 4S-AF scheme score with AF recurrence after radiofrequency ablation

|  | Univariate model |  |  | Multivariate model |  |  |
| --- | --- | --- | --- | --- | --- | --- |
|  | HR | 95% CI | P-value | aHR<sup>a</sup> | 95% CI | P-value |
| Stroke risk (St) | 2.60 | 1.36-4.97 | **0.004** | 2.14 | 0.98-4.67 | 0.056 |
| Symptoms (Sy) | 0.97 | 0.70-1.35 | 0.867 |  |  |  |
| Severity of AF burden (Sb) | 1.67 | 1.15-2.43 | **0.008** | 1.84 | 1.25-2.72 | **0.002** |
| Substrate (Su) | 1.59 | 1.35-1.86 | **<0.001** | 1.71 | 1.36-2.14 | **<0.001** |
| 4S-AF scheme score | 1.37 | 1.22-1.53 | **<0.001** | 1.38 | 1.19-1.59 | **<0.001** |
| 2S-AF scheme score | 1.53 | 1.33-1.76 | **<0.001** | 1.59 | 1.33-1.89 | **<0.001** |

AF, atrial fibrillation; HR, adjusted hazard ratio; aHR, adjusted hazard ratio; CI, confidence interval.
<sup>a</sup>Adjusted for age, gender, body mass index, coronary artery disease, diabetes mellitus, heart failure, hypertension, stroke/transient ischemic attack (TIA), atherosclerosis, estimated glomerular filtration rate.

With regard to the impact of each domain on AF recurrence, we found that the independent predictors of AF recurrence using the 4S-AF domains were Sb (aHR 1.84, 95% CI: 1.25–2.72, P=0.002) and Su (aHR 1.71, 95% CI: 1.36–2.14, P<0.001) (Table 4). The St and Sy domains did not significantly affect AF recurrence. Then, we investigated the impact of the modified scheme score combined with the Sb domain score and Su domain score on AF recurrence. Notably, multivariable Cox proportional hazards models demonstrated that the 2S-AF scheme score combined with the Sb domain score and Su domain score was a stronger independent predictor of AF recurrence (aHR 1.59, 95% CI: 1.33–1.89, P<0.001; Table 4).

### Predictive ability of 4S-AF scheme in patients who underwent RFCA

As shown in Figure 4, our ROC analysis revealed that both the 4S-AF scheme score (AUC 65.2%, 95% CI: 59.3–71.1) and the 2S-AF scheme score (AUC 66.2%, 95% CI: 60.2–72.1) had modest predictive capability for AF recurrence. Compared to HATCH (AUC 56.9%, 95% CI: 50.6-63.1) and CHA_2_DS_2_-VASc score (AUC 58.0%, 95% CI: 51.9–64.1), 4S-AF scheme score and 2S-AF scheme score had better clinical predictive capability of AF recurrence. When a 4S-AF scheme score cutoff value of 3.5 was set, patients with 4S-AF scheme score ≥ 3.5 had a higher risk of recurrent AF than those with 4S-AF scheme score < 3.5 (HR, 2.121, 95% CI, 1.449–3.106, P<0.001). Z-statistic demonstrated that there were no statistically significant differences in the AUC values between the two schemes (Z-statistic=-0.743, P=0.458). Furthermore, the AUC values of the independent predictive ability of the Sb and Su domains were 55.9% (95% CI: 49.4-62.3%) and 65.6% (95% CI: 59.6-71.7%), respectively.

**Figure 4.**
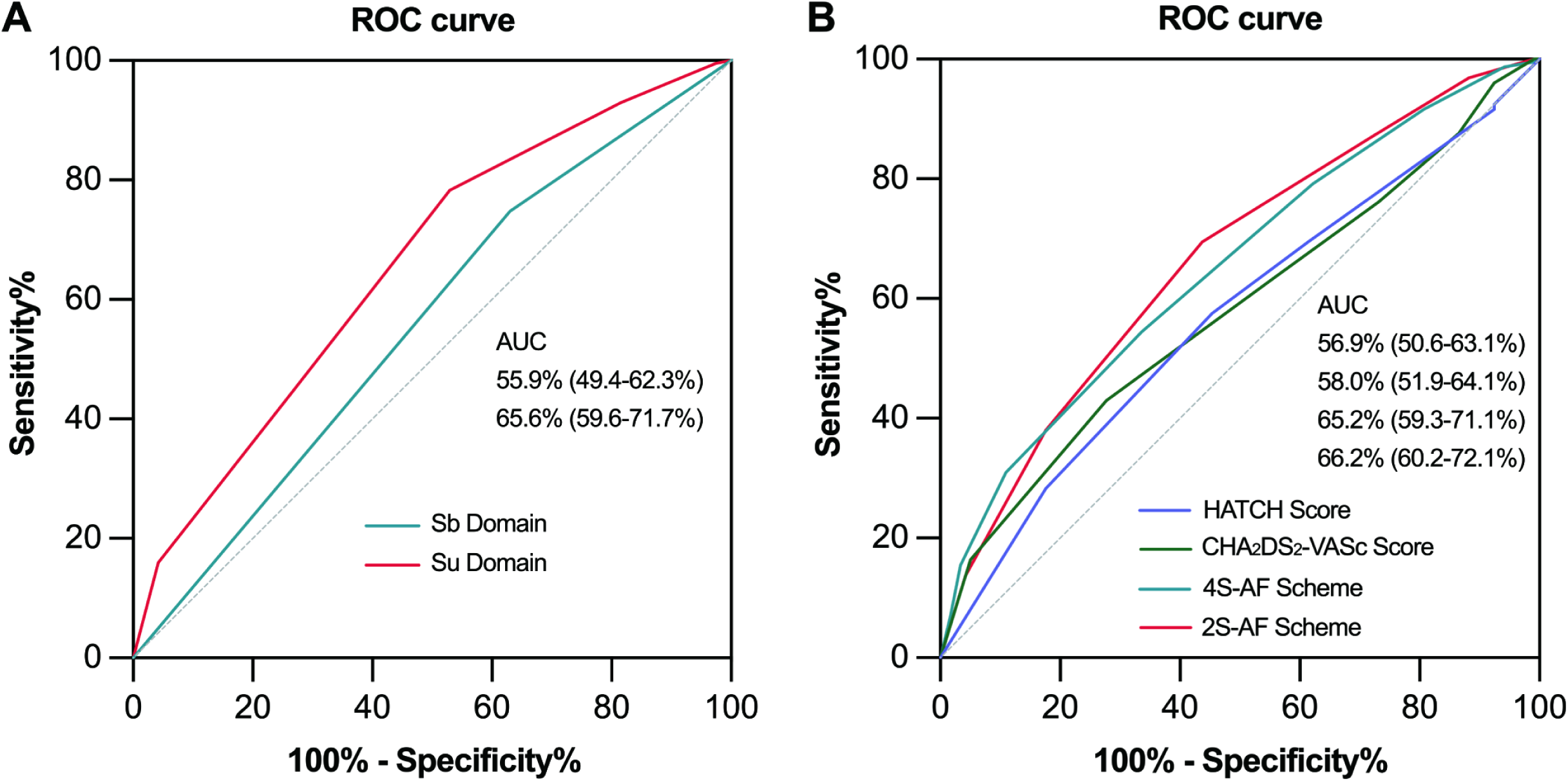
Receiver operator curve curves and corresponding AUC of Sb domain and Su domain (A); Receiver operator curve curves and corresponding AUC of 4S-AF scheme, 2S-AF scheme, CHA_2_DS_2_-VASc Score, and HATCH Score (B). AUC, area under the receiver operator curve.

## Discussion

### Principal findings

In this study, we prospectively investigated the clinical utility of the novel 4S-AF scheme in predicting AF recurrence after RFCA. The principal findings were as follows: (1) Utilizing routinely collected data, 4S-AF scheme is feasible for providing clinical characterization of patients with AF indicated to undergo RFCA; (2) a higher 4S-AF scheme score was significantly associated with an increased rate of AF recurrence after RFCA, and the 2S-AF scheme score combined with the Sy and St domains was a stronger independent predictor of AF recurrence; (3) among the 4S-AF scheme domains, Sb and Su scores were independently associated with AF recurrence outcomes after RFCA; (4) both 4S-AF scheme score and 2S-AF scheme score had modest value for predicting AF recurrence after RFCA.

### 4S-AF scheme and AF

The 4S-AF scheme reflects a paradigm shift from other classification systems towards a structured characterization of AF. This novel and practical assessment approach has been adopted and recommended in the European Society of Cardiology 2020 guidelines for the management of AF.^5^ Analysis of EORP-AF Long-Term General Registry data demonstrated that the 4S-AF scheme provided prognostic information on adverse outcomes related to AF, including all-cause mortality, CV mortality, thromboembolic events, and ischemic stroke.^26^ Notably, treatment decisions based on this scheme decreased all-cause mortality rates in patients with AF.^21, 30^ Similar findings were observed in both APHRS-AF registry cohort and FAMo cohort studies.^24, 25^ Recently, an analysis of RACE V data indicated that the 3S-AF scheme modified by eliminating the Sy domain of 4S-AF scheme may predict AF progression in the subset of self-terminating AF patients.^22^ Furthermore, the 4S-AF scheme has prognostic implications for all-cause mortality and is a pragmatic risk stratification tool for patients with new-onset atrial fibrillation after myocardial infarction.^23^ Nonetheless, studies assessing and validating the clinical utility and prognostic capability of the scheme in AF patients after RFCA are limited in number.

The 4S-AF scheme is feasible for characterizing and evaluating AF because it includes the collection of routine data on demography, comorbidities, AF-related symptoms, severity of AF burden, and LA substrate. The scheme is composed of four domains: St, Sy, Sb, and Su. St is based on CHA_2_DS_2_-VASc score, and Su domain score is composed of 3 subdomains, including the number of comorbidities/CV risk factors, atrial remodelling, and elder age. Hence, both the St domain and Su domain partly depended on comorbidities when characterizing AF patients by the 4S-AF scheme. The major limitations of this scheme are that the definitions of the comorbidities and CV risk factors remain unclear, and some conditions might be neglected during routine clinical practice. In this study, heart failure with preserved ejection fraction patients may have been underestimated due to the limited use of invasive exercise haemodynamics assessments.^31^ To evaluate atrial remodelling, we simply assessed LA enlargement by using Doppler ultrasound, which may lead to the neglect of early atrial remodelling manifesting as atrial dysfunction.^22, 32, 33^ Consequently, the total St domain score and Su domain score in the current analysis were lower than those in previous studies. For the Sy domain, the severity of symptoms was greater in the current population than those in the EORP-AF Long-Term General Registry and APHRS-AF Registry.^25, 26^ One of the reasons may be that we included symptomatic AF patients who were indicated for RFCA. Moreover, the score of the Sb domain was lower than that in other studies since we excluded long-standing persistent and permanent AF.

### 4S-AF scheme and AF recurrence after RFCA

This is the first study to use the 4S-AF scheme to characterize AF patients who underwent RFCA and to investigate its clinical utility in predicting AF recurrence after RFCA. Our analysis demonstrated that both a higher 4S-AF scheme score and 2S-AF scheme score were independently associated with an elevated AF recurrence rate after RFCA. Chollet L et al. confirmed that the combination of the AF phenotype and LAVI has prognostic value for AF recurrence after pulmonary vein isolation.^34^ Similarly, only the Sb and Su domains of the four domains in the 4S-AF scheme were found to be independent predictors of AF recurrence after multivariable adjustment in our study. Although it has been suggested that the 4S-AF scheme is not to be used for the purpose of risk stratification, in this prospective cohort, we validated that the 4S-AF and 2S-AF schemes had modest predictive performance for post-RFCA AF recurrence. These findings suggested that using this scheme to characterize AF may aid in identifying which high-risk patients should undergo RFCA for rhythm control. However, data from the EORP-AF Long-Term General Registry indicated that higher 4S-scheme scores were suitable for more aggressive interventions to improve the adverse long-term outcomes of AF.^26^ Hence, the use of the 4S-AF scheme to predict different post-RFCA outcomes and the weighting of each domain need future validation to better streamline the process of selecting AF patients who may benefit from RFCA.

Although only a modest predictive performance of the 4S-AF scheme was found in this study, the clinical utility and prognostic value of the 4S-AF scheme and its modified schemes for predicting AF recurrence after RFCA are highly desirable for further investigation. Numerous emerging studies have validated that several novel biomarkers, such as electrocardiographic, molecular, and imaging biomarkers, can independently predict AF recurrence outcomes after RFCA.^35–40^ In combination with these novel biomarkers and other well-established scoring systems, the predictive ability and reclassification performance of the 4S-AF scheme score may be significantly improved among patients who undergo RFCA.^41^ Given that identifying atrial remodelling and monitoring cardiac rhythm by routine tools in clinical practice are challenging, most descriptors of AF domains have yet to be defined appropriately and evaluated accurately. We believe that the 4S-AF scheme has great clinical utility and prognostic value with future refinements guided by advanced cardiac imaging and rhythm screening technology.^42^

### Study limitations

We acknowledge several limitations of the current study. First, we included only paroxysmal and persistent AF patients who underwent initial RFCA at our centre in this analysis, and the exclusion of a significant proportion of patients may be a source of bias. Therefore, the conclusions should be validated by future studies in more diverse cohorts of AF patients after RFCA. Moreover, the tools available to assess AF episodes and burden, including 24-h Holter monitoring and 12-lead ECG, may lead to underestimation of AF recurrence compared with implanted or continuous rhythm monitoring devices.^22, 43^ The 4S-AF scheme is a dynamic score, but we only assessed it at baseline and had limited follow-up data for periodic reassessment. Furthermore, there is some overlap between the four domains. Finally, this was a single-centre with a limited sample size. AF is a complex condition, and we cannot exclude residual confounding.^3^

## Conclusions

We demonstrated that the novel 4S-AF scheme is feasible and practical for evaluating and characterizing AF patients who undergo RFCA. Characterization using this novel scheme independently assisted us in identifying who was at high risk of AF recurrence and predicting AF recurrence after RFCA.

## Data availability

The data underlying this article will be shared on reasonable request to the corresponding author.

